# Exploring potential of AI usage in the knowledge and evidence services of a public health body: a working group approach

**DOI:** 10.1101/2024.07.08.24310046

**Authors:** Zalaya Simmons, Charlotte Bruce, Samuel Thomas, Patricia Lacey, Wendy Marsh, Scott Rosenberg, Daphne Duval

## Abstract

The UK Health Security Agency (UKHSA)’s Knowledge and Library Services (KLS) established an Artificial Intelligence (AI) working group in 2022 to explore potential applications of AI of relevance to its function. This paper describes the working group’s approach to testing and evaluating AI and machine learning-assisted tools for information retrieval and evidence review processes, including duplicate reference removal, citation searching, title and abstract screening, full text screening, data extraction and critical appraisal. Initial tests have demonstrated varying degrees of potential for implementation, while also contributing to broader discussions on ethical considerations, copyright and licensing issues, transparency of AI methodology and evidence integrity. This overview outlines the methodology used and insights gained from navigating the rapidly evolving AI landscape and its potential implications for knowledge and library services within a public health organisation.

## Introduction

The UK Health Security Agency (UKHSA), an executive agency of the Department of Health and Social Care (DHSC), aims to prevent, prepare for and respond to infectious diseases, and environmental hazards. It also provides scientific and operational leadership, working with local, national and international partners to protect the public’s health and build the nation’s health security capability. The Knowledge and Library Services (KLS) within UKHSA facilitates access to evidence through a range of mediated, embedded library, information and knowledge services delivered at the point of need, including expert literature searching, information retrieval, evidence reviews, document delivery and inter-library loans, access to resources (journals, databases, etc) and knowledge management services (UKHSA, 2023a). KLS comprises of 35 members of staff organised into different teams: knowledge and evidence specialists, knowledge management and mobilisation team, evidence review team, and library operations team (system librarians and on-site librarians).

In this paper, we mainly focus on two of the core services: literature searching and evidence reviews. For literature searching, KLS offers several search output levels that include a simple bibliography, results categorised by theme, evidence summaries and searches for systematic reviews. Since 2023, the offer has been extended to evidence synthesis outputs (especially rapid reviews, scoping reviews and mapping reviews) which all follow systematic methodologies: protocol development, database searches, title and abstract screening, full text screening, data extraction, critical appraisal and evidence synthesis.

Not all KLS staff undertake evidence reviews, literature searching or produce the same outputs (depending on job role and user group). Within the KLS team, there are currently 4 evidence reviewers who conduct evidence reviews (but no searches) and 17 KLS staff who regularly undertake literature searches as part of their day-to-day work, although not all 17 produce all types of search outputs (for instance, not everyone conduct searches for systematic reviews).

In 2022, an AI working group was created within KLS to conduct horizon scanning and explore the rapid advancement and increasing availability of Artificial Intelligence (AI) tools, of potential application to KLS services, especially literature searching and evidence synthesis. In this article, we provide an overview of the work undertaken in KLS in relation to AI, from developing and establishing an AI working group, to the testing of AI tools within the literature searching, screening and data extraction processes. Not all tools tested have been implemented within KLS services, but learning from all testing has been used to further inform AI working group activities.

## The KLS AI Working Group

The KLS AI working group was set up in July 2022 by members of staff within KLS with an interest in AI topics. This was in response to the increase in research on AI being presented and published, including the 2021 CILIP report ‘*The impact of AI, machine learning, automation and robotics on the information professions’ (Cox, 2021)*. Following an evaluation of possible options on how to engage with AI within KLS, it was determined that a designated working group, chaired by members of the KLS team, would be the most suitable option to explore AI and emerging technologies.

The initial terms of reference were developed and written by the first chair of the group in collaboration with KLS colleagues and approved by our KLS senior management team. The working group meets monthly (virtual meeting via Microsoft Teams) and attendance is open to anyone within KLS. When created in 2022, the working group had 10 members; it currently has 14 members (as of June 2024).

The working group focuses on how AI and emerging technologies could be used within KLS and has included software testing and awareness raising, but also supporting the development of the wider health librarianship community by sharing the work of the group externally. In the first months, the working group shared learning on AI technologies gained at conference and meeting attendance and started developing its approach to testing. It was agreed that the working group would focus on testing specific AI based emerging technologies using a collaborative approach to monitor and record testing outcomes using Microsoft Teams. The initial chair of the working group assigned group members to lead the testing of specific software to develop a distributed leadership model within the group. This proved effective in fostering a collective sense of engagement, collaboration and concurrent leadership within the working group (Chilton, 2004).

As the working group has progressed, it has become a forum for members to exchange ideas and conduct horizon scanning of AI and emerging technologies. Pertinent topics discussed have included user enquiries received by KLS on using AI, generative AI, ethics and governance, prompt engineering, copyright considerations and evaluation of AI software. Since its creation there have been many successful outcomes, including:

- introduction of new technologies into literature searching and the systematic review process
- mobilisation of knowledge on AI topics among the KLS team, increasing awareness and confidence
- invitation of guest speakers from a variety of organisations working with evolving AI tools to share learning around use of AI
- promotion of the work of the group to a wider audience, for example, group members presented to the NHS Futures AI community and at the 2022 NICE Joint Information Day, facilitating knowledge sharing and networking between organisations

The AI working group and KLS team have explored and tested various AI technologies and software to assist activities the KLS team routinely undertake. Notably this has been within literature searching, screening of search results and the data extraction stage of the review process, as areas in which new technologies can potentially assist in the work of a significant proportion of KLS staff.

## Literature searching

The working group has explored several AI technologies to support information retrieval processes, including Citationchaser (citation searching) and Deduklick (duplicate reference removal in search results). These tools were tested formally (using predetermined criteria, multiple testers, and comparisons to similar technology) or semi-formally (testing conducted over a shorter timescale with singular testers) and both are now routinely used by KLS staff. In the following, we describe more details of the testing conducted for each of these tools.

### Citationchaser

Citation searching is a supplementary search method that identifies relevant documents by utilising explicit references made from one article to another (Haddaway et al., 2022). Citation searching can be ‘backward’ finding referenced articles in a bibliography or reference list, or ‘forward’ finding articles in which the reference is included within. Citationchaser automates this process using R and open source software in an “easy-to-use tool” via a user-friendly graphical interface (Haddaway et al., 2022). Citation searching can be a time-consuming search method and is undertaken to differing degrees and frequency by KLS staff, most common in searches for evidence briefings and evidence reviews.

Due to the potential benefits of Citationchaser, the software was tested in November 2022 by a member of the working group who regularly undertakes citation searching. The aim of this testing was to explore functionality via semi-formal testing structured into three stages (using search results from one literature search undertaken by the tester in that same month):

1. Overall functionality and usage, using 18 references (articles and grey literature, reports)
2. A comparison of three references (articles) with Scopus (a database previously used by the tester for citation searching) to explore similarities and differences.
3. A singular reference, retrieved by both Citationchaser and Scopus, forward citation references retrieved compared.

A summary of these testing stages is presented below.

#### Stage 1 – Overall functionality and usability

To test functionality and ease of use, 18 references were selected from an existing EndNote library of search results. The references were separated out into two groups as it had been hypothesised that the grey literature references would not be identified by Citationchaser: group 1 included 13 journal articles (11 retrieved from bibliographic database searches, two manually added into EndNote by the searcher) and group 2 included five grey literature references with reference type ‘Report’ (all five were manually added by the searcher to EndNote). The two groups were tested separately in Citationchaser, uploaded via RIS in a text file format, for forward and backward citation searching.

Out of the 18 references tested, Citationchaser was only able to identify the 11 references that had been imported from bibliographic databases. With these 11 references, it was able to locate (using both forward and backward citation searching) 399 unique references (backward citation) and 699 unique references (forward citation searching). This showed that rather than the reference type, issues arose from how the references were imported (retrieved from database versus added manually).

Whilst some metadata was missing (such as DOI or abstract) from the references retrieved by Citationchaser, most references contained enough information to be reviewed. References retrieved included some foreign language articles and a very limited number from grey literature. Overall, Citationchaser was found to be simple to use from upload to downloading results and was a fast way to undertake citation searching of multiple references.

#### Stage 2 – Scopus comparison

From the 11 references retrieved from bibliographic database searches, three were chosen at random to compare results between Citationchaser and Scopus for single citation searching (backward and forward). Results are shown in Table 1.

**Table 1.** Number of references identified for backward and forward citation searching by Citationchaser compared to Scopus for the 3 references tested.

|  | Backward citation searching |  | Forward citation searching |  |
| --- | --- | --- | --- | --- |
| Reference number | Citationchaser | Scopus | Citationchaser | Scopus |
| 1 | 84 | 84 | 377 | 325 |
| 2 | 30 | 31* | 67 | 46 |
| 3 | 23 | 34* | 10 | 7 |

\*shows the correct number of references, based on article bibliography

There was a difference in numbers retrieved in all but one instance of forward and backward citation searching (reference 1 backward citation searching). Scopus was able to provide the most accurate backward citation searching and give basic information on references not indexed in its database, using the bibliography details (of the reference used for citation searching), whereas Citationchaser could not. This was most evident with bibliographies with many grey literature references, identifying a limitation of Citationchaser.

Comparatively, Citationchaser retrieved more references than Scopus in forward citation searching for all three test references. This may be because Citationchaser is using more sources than Scopus, including the Lens.org database (which consists of PubMed, PubMed Central, CrossRef, Microsoft Academic Graph and CORE) (Haddaway et al., 2022).

#### Stage 3 – Reference comparison

Due to the differences observed in stage 2 testing for forward citation searching, one reference (reference 3) was selected for more detailed comparison on forward citation searching results. Both sets of forward references were imported into EndNote and compared to see common and unique references. Only four references (8 when including the duplication) were retrieved by both Citationchaser and Scopus. Citationchaser retrieved 6 unique references compared to Scopus’ 3 unique references. These results suggest that Citationchaser is a valuable tool for fast citation searching of multiple references, however, it is also evident that there is value in using more than one citation searching platform due to the number of unique references each tool identified.

#### Adoption of Citationchaser

The testing of Citationchaser provided valuable information on the benefits and potential limitations of the technology before suggesting adoption in the KLS search process. Based on this testing, the members of our working group were encouraged to use the application and share their experiences. However, it is also important to note that the testing undertaken was semi-formal and conducted by only one member of the working group. In particular, it would have been valuable to do further comparison, such as reviewing more results, exploring different search topics, and with other databases such as Web of Science.

In June 2024, circa 18 months after Citationchaser was introduced in the team, a member from the working group conducted a survey (10 respondents, of whom 80% used Citationchaser) and collected informal feedback from the KLS team to gain insights about its use within the team. Results showed that many of the KLS staff use Citationchaser in combination with other platforms or sources (such as Web of Science, Google Scholar and manually checking reference lists). KLS staff reported that Citationchaser saves time and encourages greater citation searching, but that when using it they may limit the number of references they import (due to number of results retrieved).

It was found that Citationchaser is now routinely used by KLS staff undertaking literature searches, although it has not been universally adopted. Adoption has however been seen more formally in some processes and outputs within KLS, for example in rapid reviews for which citation searching was not usually undertaken due to time limitations.

### Deduklick

Literature searches are usually conducted across multiple databases in order to ensure a robust interrogation of the evidence-base and reduce the risk of omitting relevant literature (Ewald et al., 2022). However, as there is some overlap between the databases in terms of indexed journals, it is important to identify and remove duplicates. KLS staff used to do this in EndNote (the reference management software used by KLS) using the Leeds University Library methodology (Falconer, 2018), referred to in this paper as the Leeds Method. This method is effective but time-consuming, requiring KLS staff to change settings within EndNote multiple times. In this context, the working group identified Deduklick, an AI duplicate identification and removal software which has been found to have high precision in finding unique references and saves time (Borissov et al., 2022) as a possible alternative to the Leeds method.

Deduklick was formally tested in June and July 2022 whilst undertaking a trial of the product. Four testers within the working group and wider KLS team compared Deduklick and the Leeds Method on eight literature searches they were undertaking at that time. A lead tester was assigned to organise the testing and collate the data retrieved. Data was collected in Excel and included the search topic, the type of search output (based on KLS service offer), databases used, import order into EndNote and deduplication numbers for each method. Searches used for testing were on a variety of public health topics and were used for a range of search outputs. There were 15 unique databases used across all searches, and the number of databases per search varied (from three to seven). The number of search results (prior to duplicate removal) also varied considerably, ranging from 253 to 8,986. The number of identified duplicates from each method is displayed for each search in Figure 1.

**Figure 1.**
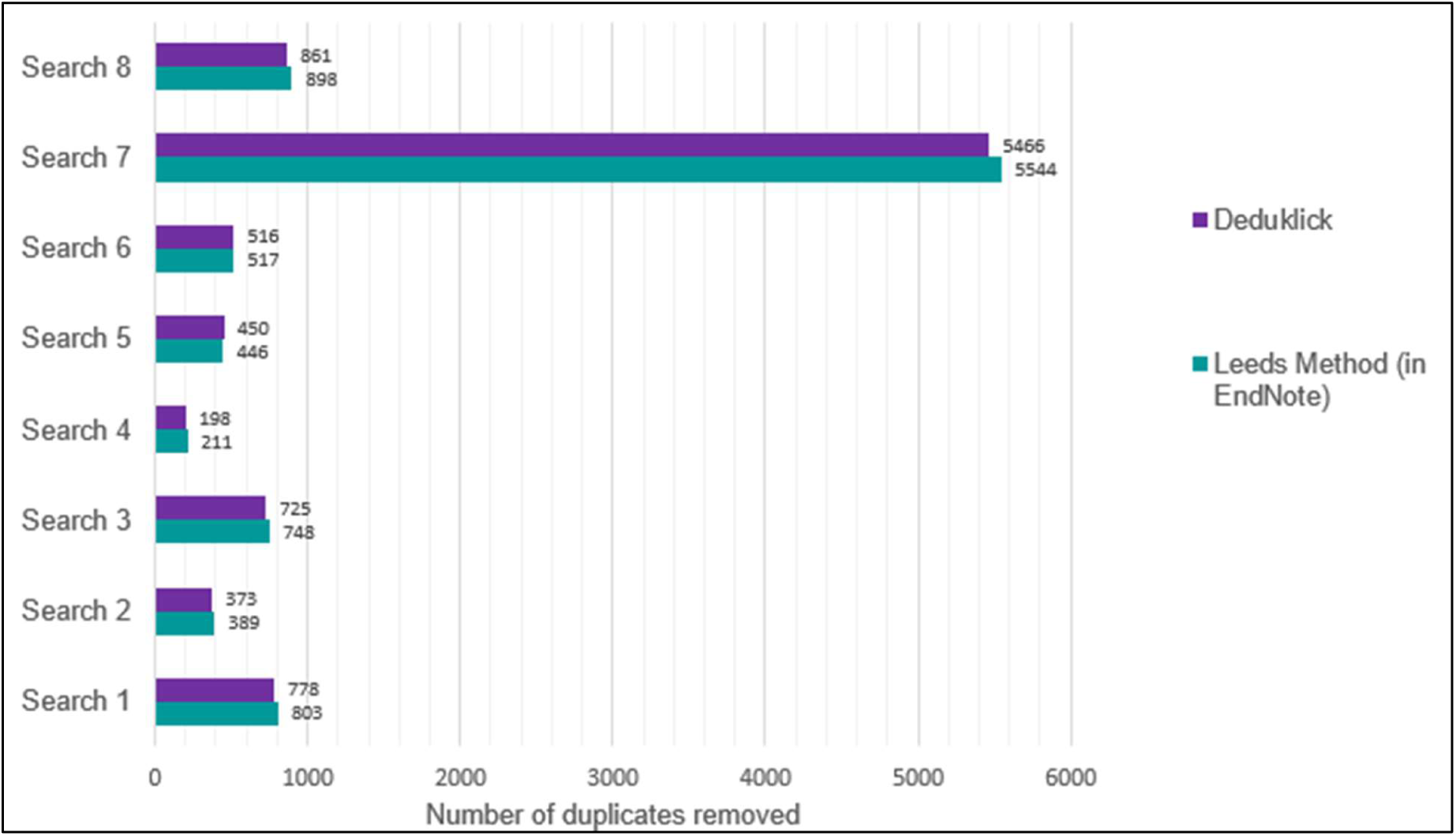
Number of duplicates removed using Deduklick and Leeds method.

Results showed that Deduklick removed more references than the Leeds Methods in two searches: differences in identified duplicates of 0.9% for Search 5 (450 versus 446) and 4.2% for Search 2 (373 versus 389). However, Deduklick identified less duplicates than the Leeds Method in the six other searches (Searches 1, 3, 4, 6, 7, and 8), although the differences in identified duplicates ranged from 0.2% for Search 6 (516 versus 517 duplicates) to 6.4% for Search 4 (198 versus 211 duplicates), which was judged acceptable by the working group.

Based on these findings, the working group concluded that Deduklick performed similarly to the Leeds Method while saving considerable staff time. As a result, the software was purchased in April 2023.

As KLS staff have become more experienced in using the technology they have noted that Deduklick retained results which were actual duplicates but not exact matches (for example, differences in record metadata) and that the Leeds Method would have correctly identified them as duplicates. However, the benefits of using Deduklick, including staff time saved, and the lower risk of error (removing non exact matches) was considered satisfactory when compared with its overall lower number of duplicates removed.

Deduklick has been widely adopted by the KLS team: from purchase in April 2023 to June 2024 there are currently 19 registered users. We reviewed Deduklick user accounts in June 2024 and categorised them by number of unique searches and role of the user within KLS. This showed that 13 accounts belong to active searchers in KLS (out of 17 KLS staff who regularly undertake literature searches as part of their day-to-day work, this is, a 77% uptake). However, usage numbers vary as half of accounts (10 total) sit within a low usage banding (between 0-20 unique uses), with remaining accounts in medium usage (between 21-50 unique uses; 4 accounts) and high usage (over 51 unique uses; 3 accounts). The variation in usage is influenced by numerous factors. In particular, the three high usage accounts are all staff undertaking searches for systematic reviews, which are not undertaken by all staff. In addition, frequency of searching varies among staff, and staff may decide not to use Deduklick for every search, for example if there are a small number of results retrieved.

Finally, although usage has increased over the last fifteen months, partly due to internal promotion of the benefits of the technology, not all KLS staff who regularly undertake literature searches currently use Deduklick. When informally asked why, reasons cited include needing the time to learn to use Deduklick (but do intend to use in the future), feeling happy with the current duplicate removal methods used and preferring to have control over which record is retained or removed.

## Title and abstract screening

In a systematic review, title and abstract is done in duplicate (two reviewers screening the same dataset independently), whilst in a rapid review only a percentage of the records to assess are screened in duplicate (typically 10 or 20%) and the remaining is screened by only one reviewer.

Commercial machine learning tools are available to help accelerate the title and abstract screening process (dos Reis et al., 2023) and research in this area suggests they can be effective at accelerating the screening process for evidence synthesis (Burgard and Bittermann, 2023). These tools use various methods, such as active learning and machine learning classifiers, to identify potentially relevant references and prioritise them to the top of the screening list, while pushing potentially irrelevant references to the bottom. In 2023, the working group began exploring the use of these tools for screening literature search results for various outputs, such as evidence briefings and rapid reviews. In the following, we present the results of the testing of two commercially available screening tools which use machine learning assisted title and abstract screening: Rayyan and EPPI-reviewer.

### Rayyan machine learning-assisted screening (title and abstract)

Rayyan is a web-based review management tool that facilitates reference management and the screening process for evidence reviews (Mourad Ouzzani, 2016). They offer a free to access and subscription-based package, both of which include a built-in machine learning classifier, which uses a Support Vector Machine (SVM) classifier to learn from users’ inclusion and exclusion decisions and prioritise relevant references to the top of a list for screening. The classifier uses features such as unigrams, bigrams, and MeSH terms extracted from the title and abstract of each study which are used to train the SVM classifier based on the reviewers’ initial screening decisions (that is, labelling citations with an include or exclude decision). The model then provides citations with a relevance score, represented as a five-point rating. As more citations are screened by the reviewers, the model is updated iteratively, and more precise recommendations are provided for the studies that are still awaiting screening.

The Rayyan prediction classifier was first tested informally in 2023 by a member of the AI working group, showing promising results (typically, all relevant records were found in the first 30/40% of the prioritised list). However, the use of the Rayyan prediction classifier had not been yet formally implemented in KLS review process due to the lack of formal testing. In this context, an evidence reviewer from KLS tested the Rayyan prediction classifier more formally in June 2024, using data from a rapid review conducted by the team in early 2024. For this review, 2,581 deduplicated references had been screened on title and abstract (10% in duplicate, the rest by one reviewer; as per our rapid review processes), of which 66 references were included for full text screening. Full text screening was done by one reviewer and checked by a second, and 16 references were included in the final review.

To evaluate the effectiveness of Rayyan’s prediction classifier, the title and abstract screening was replicated using the same set of 2,581 deduplicated references. Two reviewers screened 10% of the references in duplicate, of which 12 were included on title and abstract. These initial screening decisions were then used as training data, and Rayyan ratings were computed. One reviewer (who had not been involved in the original screening) continued to screen the remaining references (n=2,320) and prompted Rayyan to recalibrate its ratings every 50 references screened, until all references had been screened. In total, 58 records were included on title and abstract: 12 in the training data, and 46 once the Rayyan ratings had been computed.

The number of records included on title and abstract in function of the number of records screened once Rayyan ratings had been activated are presented in Figure 2. This graph shows that records included on title and abstract were found early on in the screening process before reaching a plateau, which suggested that relevant papers were found at the top of the prioritised list. More concretely, 96% of references included once ratings had been computed were found within the first 13% of the prioritised list (that is, 44 out of the 46 references included after ratings were computed were located within the first 300 references of the prioritised list), and all 100% studies were found within the first 32% of the prioritised list (46, located within the first 750 references of the prioritised list).

**Figure 2.**
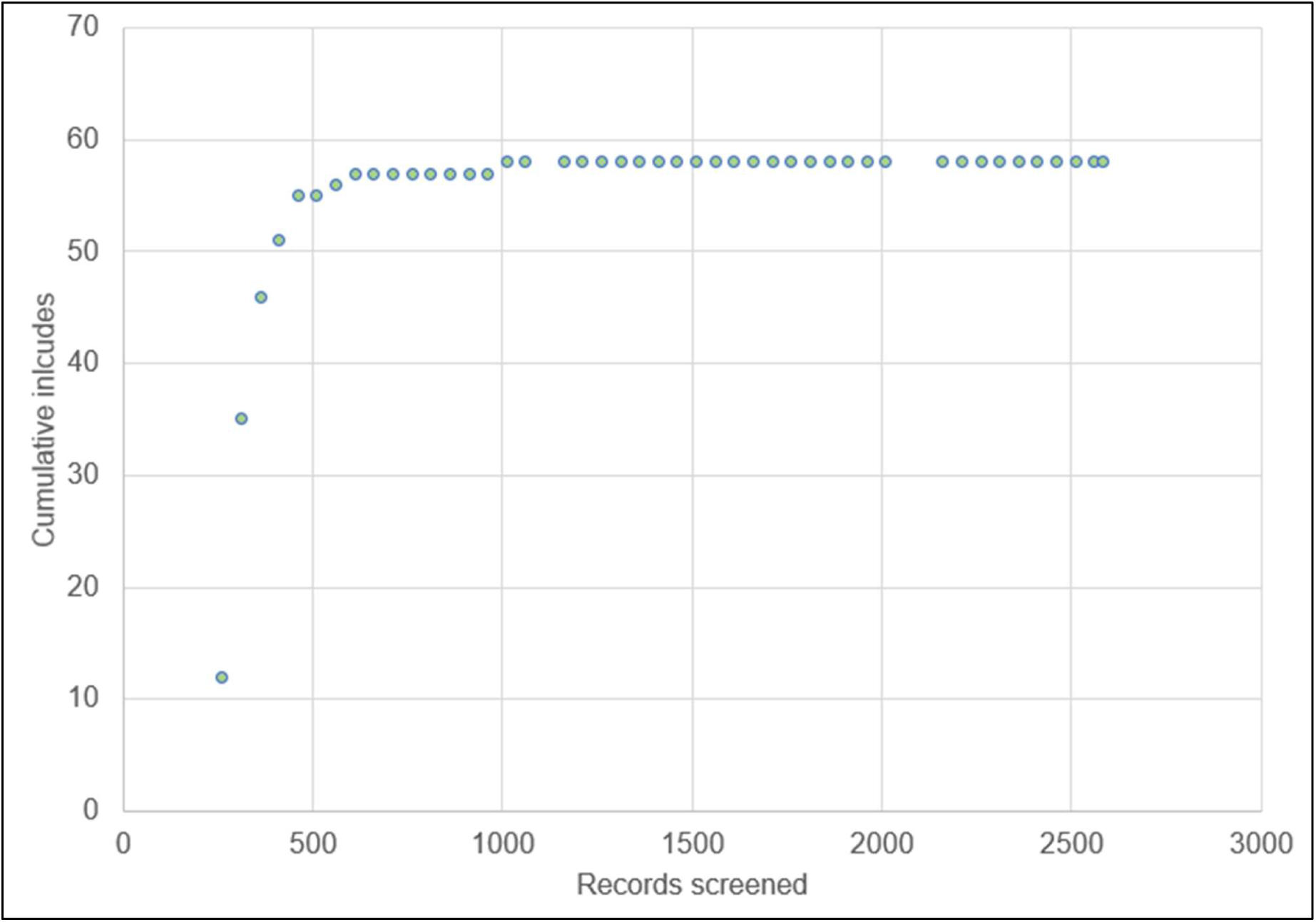
Number of records included on title and abstract in function of the number of records screened using Rayyan ratings (machine learning-assisted screening)

To assess the effectiveness of the Rayyan prediction classifier in identifying the most relevant studies, we retrospectively analysed at what position within the prioritised list the 16 references included in the final review first appeared. Notably, all references included in the final review were identified within the first 150 records screened after prioritisation, accounting for just 6.3% of the 2,320 references screened after prioritisation was initiated. This demonstrates how effective Rayyan was at identifying relevant references early in the screening process despite a relatively small amount of training data (261 references, of which 12 had been labelled as “include”).

**Figure 3.**
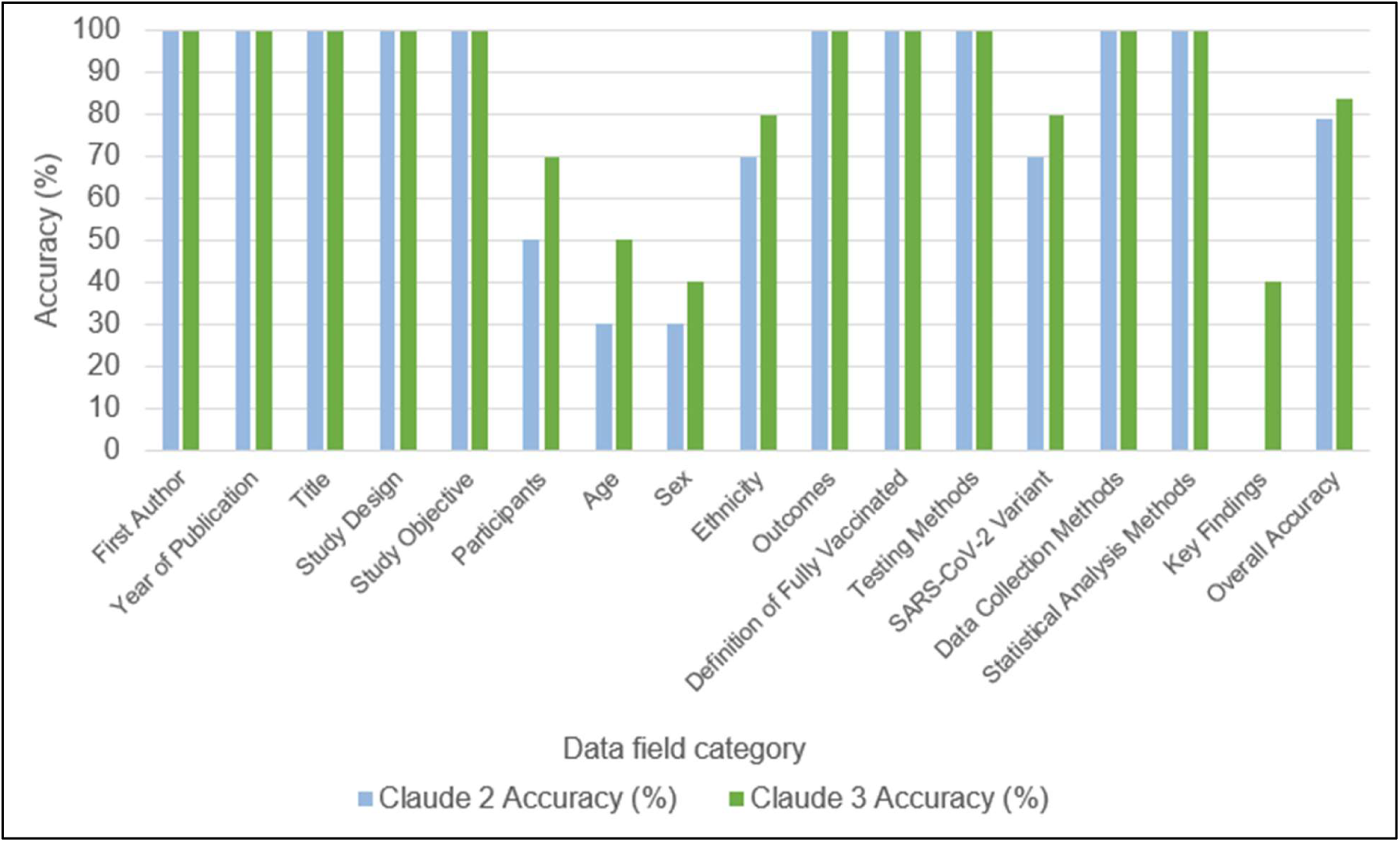
Comparison of accuracy between Claude 2 and Claude 3 across data field categories for data extraction.

### EPPI-reviewer machine learning-assisted screening (title and abstract)

EPPI-Reviewer (EPPI-R) is another systematic review management tool that offers a machine learning-assisted screening function (EPPI Reviewer priority screening), which is designed to accelerate the screening process using machine learning (Thomas, 2023). Similarly to Rayyan, the EPPI’s priority screening function prioritises references by predicting their relevance based on initial training data provided by users’ screening decisions. In 2023, evidence reviewers from KLS tested this function while completing title and abstract screening for a rapid review. A literature search was conducted and, after deduplication, 15,427 references were imported into EPPI-R. As with the Rayyan ratings study, the machine learning model was trained on human screening decisions, representing 10% of the total references (1,543 references) which were screened in duplicate as per our rapid review methods.

Once trained, the model re-ordered the remaining 13,884 references, bringing potentially relevant studies to the top of the list. This re-prioritisation occurred at intervals, starting every 25 references and less frequently as more references were screened. During our testing, we found that 95% of the studies included on title and abstract were identified within the first 25% of the prioritised list (that is, 122 out of the 129 records included on title and abstract screening were located within the first 3,477 references), and 99% within the first 30%. These results demonstrate the tool’s capability to prioritise potentially relevant studies effectively.

### Potential for implementation of machine learning-assisted screening (title and abstract)

The findings from these evaluations demonstrate the potential of machine learning tools to enhance the efficiency of title and abstract screening for evidence synthesis. Both tools were effective at prioritising potentially relevant papers, accelerating the overall review delivery. In a rapid review, machine learning-assisted screening enables parallel working as once the plateau is reached (that is, when most of the records are being screened as excluded, see Figure 2), one reviewer can start full-text screening to identify relevant references rapidly while the other reviewer continue title and abstract screening until all remaining references had been screened. In a systematic review, the machine learning-assisted screening can act as one reviewer, with a human reviewer also screening all references, therefore acting as a second reviewer for duplicate screening. These approaches result in more efficient processes (by saving time or/and needed less reviewers) while not compromising on screening accuracy as all references are still screened by one human reviewer.

Machine learning-assisted screening (title and abstract) using both Rayyan and EPPI-R is now used regularly within KLS services, both for evidence summaries and rapid reviews, although it has not been universally adopted. Indeed, the decision to use machine learning-assisted screening is done on a case-by-case basis, dependent on factors such as the complexity of review inclusion criteria, the number of records to screen and the number of reviewers involved in the screening process. Further testing of these tools is required to ensure reliability and effectiveness across a wider range of review questions, and continuous monitoring to identify any biases that may arise from the machine learning algorithms.

Moving forward, to further improve the working group’s understanding of machine learning options and their capabilities, limitations and applications for title and abstract screening, the working group plans to explore other machine learning methods such as text mining in future testing. Additionally, we will evaluate these tools on more diverse review questions, including reviews with more complex inclusion criteria and diversity of study designs and publication types.

## Full text screening

Full text screening, the second stage of the screening process for evidence reviews, involves assessing the complete texts of studies that were included on title and abstract. In a systematic review, full text screening is undertaken in duplicate (two reviewers screening the same dataset independently). Different shortcuts can be used for full text screening in rapid review: one approach, similar to the one used for title and abstract screening, consists in screening only a percentage of the records in duplicate. The other approach, which is used by the KLS evidence review team, is to have a second reviewer checking all the full-text that had been excluded to ensure no relevant paper had been missed.

While fewer studies are typically screened at this stage, the requirement to thoroughly evaluate the entire study text against each inclusion criteria of the review can be time-consuming. To address this, in 2023, a member of the working group began exploring the feasibility of using generative AI tools such as Claude 2 for full text screening, with a second human reviewer verifying all inclusion decisions made by the model.

Informal testing of Claude 2 was conducted to evaluate its capability to accurately apply predefined inclusion criteria to the full texts of primary studies. This involved using a previously conducted rapid review as test data, providing Claude 2 with a comprehensive prompt (which included task instructions, the review question and the review inclusion criteria), and 16 open access full study texts for assessment. In this testing, true positives and true negatives were classified as instances where both the AI and human reviewers agreed on inclusion and exclusion decisions. False positives were instances where the AI included a study that the human excluded, and false negatives were instances where the AI excluded a study that the human included. Based on these classifications, it was found that Claude 2 achieved an overall accuracy of 88%, with human reviewer decisions and Claude 2 decisions in agreement for 14 out of 16 studies screened. A misapplication of the outcome criteria was the cause of the conflict in inclusion decisions for 2 studies. However, in both cases of conflict, Claude 2 provided a reasonable justification for its inclusion decision, and they were borderline cases in which a human reviewer could have made similar arguments. Conflicts around outcome criteria demonstrated where further clarification of inclusion criteria might be necessary, or where subjective interpretations of study relevance and outcomes can contribute to conflicting decisions.

Overall, while the sample of studies screened was small, this informal testing suggests that generative AI tools like Claude 2 may have some potential to help accelerate full text screening and reduce the workload of human reviewers by screening in the place of a first reviewer, while maintaining relatively high standards of accuracy through secondary human checking of all decisions. Further large-scale testing and validation will be necessary before considering the implementation of these tools into the evidence review workflow. Importantly, a limitation of using externally hosted large language models such as Claude 2 for full text screening, is the inability to upload subscription-based full texts due to copyright and licensing restrictions, and due to this limitation, the working group is not actively testing externally hosted large language models for full text screening.

## Data extraction

Alongside literature searching and screening, data extraction is another review stage for which there may be potential for AI to save time and resources. As per previous steps of the review process, data extraction is carried out in duplicate in a systematic review. In a rapid review, the KLS approach is to have a second reviewer checking the data extraction of another reviewer.

Currently, no widely adopted extraction AI assisted tools exist that fully automate the data extraction process; however, work in this space is progressing rapidly (Gartlehner et al., 2023; Sun et al., 2024). In 2023, a member of the working group began scoping the feasibility of using large language models to assist with data extraction as a first reviewer, with a human second reviewer verifying all extractions. Using the same previously conducted rapid review used for full text screening testing, Claude 2 was tasked with extracting 16 data fields previously extracted by a human reviewer. For this informal testing, Claude 2 was provided with a prompt which included task instructions, the data fields to be extracted, and an example of the required detail and editorial style for data extraction outputted, along with the PDFs of 10 open access studies to be extracted. The testing was repeated in 2024 under the same conditions with the newest Claude 3 Opus model.

To measure the model’s performance in data extraction, accuracy, precision and recall were calculated. Accuracy provides an overall sense of the model’s performance by measuring the proportion of correct extractions (both true positives and true negatives) out of the total extraction attempts. True negatives in this context refer to the model correctly identifying data that should not be extracted, although this is less applicable since data extraction focuses on identifying relevant information. Precision measures the proportion of correctly extracted relevant data fields (true positives) out of all data fields identified as relevant by the model (true positives and false positives). Recall evaluates the model’s ability to correctly identify all relevant data fields that should have been extracted according to the human reviewer (true positives and false negatives), this is important for ensuring that significant data is not missed. Together, these metrics help to give an understanding of how effectively the model identifies and extracts relevant data, and its completeness in identifying all necessary data compared to a human reviewer’s standards.

For the ten studies that underwent data extraction by Claude 2, overall accuracy was 79% across the 16 data fields extracted for each study.

Claude 2 demonstrated high accuracy for more straightforward data fields such as first author’, ‘year of publication’, ‘title’, ‘study objective’, ‘statistical analysis’, and ‘data collection methods’, achieving 100% accuracy across these fields. However, for more complex data fields, such as ‘participants’ and ‘key findings’, accuracy was lower, at 50% and 0% respectively. While Claude 2 did extract the key findings reported in the studies, it consistently focused on the studies’ own reported key findings, rather than identifying and extracting the findings most relevant to our review question. Similarly, Claude 2 often extracted the overall participant figures, rather than identifying groups of participants relevant to the review question. Repeat testing with Claude 3 showed improvements, with overall accuracy rising from 79% to 84%. For more complex extractions, accuracy increased from 50% to 70% for ‘participants’ and 0% to 40% for ‘key findings’, with better identification of relevant data for both data fields, in particular data that was reported in tables.

Both models faced challenges with fields requiring contextual judgment and data extraction from tables. For instance, they struggled to extract subjectively relevant key findings such as secondary outcomes or subgroup analyses that were relevant to the review question. Extracting participant demographic data such as age, sex, and ethnicity, which were often reported in tables or supplementary files, also proved challenging. Despite these limitations, generative AI tools like Claude 2 and Claude 3 show potential to partially automate data extraction in evidence reviews in place of a first reviewer, reducing the workload of human reviewers while maintaining accuracy with human quality assurance. As large language models evolve, formal testing on larger, more diverse datasets will be necessary to further assess their utility and reliability for data extraction in evidence reviews. However, the inability to upload subscription-based full texts due to copyright and licensing restrictions remains the main limitation of using externally hosted large language models.

## Limitations of AI in literature searching and evidence synthesis applications

While the working group’s initial testing of AI for some information retrieval and evidence review tasks such as screening and data extraction have shown promise, testing for more subjective and complex tasks has demonstrated significant limitations. One such example is critical appraisal of study quality and risk of bias, which is a fundamental process conducted by knowledge and evidence specialists and evidence reviewers throughout KLS. This process is important for assessing methodological quality and identifying potential biases in studies and is an important methodological task for ensuring the integrity and transparency of the evidence we produce (Tod et al., 2022).

Members of the working group have conducted informal testing using large language models like ChatGPT 4.0 and Claude 2 to investigate their ability to perform critical appraisal of studies. These models were instructed to systematically evaluate studies across various risk of bias and methodological domains. Specifically, tasks included providing justified decisions for each risk of bias criterion judged, supported by examples from the study text, and assigning an overall quality or risk of bias grade using a specific appraisal framework. Regardless of the appraisal framework used or studies provided, the critical appraisals conducted by the models consistently lacked the necessary depth and discernment required for a comprehensive assessment. While they could identify text relevant to specific appraisal domains, they failed to move beyond term matching, to performing a meaningful analysis of study quality or risk of bias. Similarly, informal testing conducted so far for narrative synthesis have not been conclusive.

Tools such as Scopus AI, Dimensions (Hook et al., 2018), Semantic Scholar, Scite (Nicholson et al., 2021) and Connected Papers (Behera et al., 2023) are developing rapidly in this space, using methods like retrieval-augmented generation, natural language processing, machine learning and semantic search. Whilst useful to get a quick overview of a topic, the tests conducted so far do not suggest that these tools can meet UKHSA’s needs at this time to summarise evidence for evidence synthesis application. The working group’s ongoing horizon scanning of these technologies will help position KLS to not only make judgements on if and how they may be integrated into its services, but also educate users on their benefits, limitations and potential risks.

While informal, these results highlight the current limitations of using AI for tasks that require subjective analysis and understanding of context, and reaffirm that while powerful, large language models, machine learning and other AI subtypes are probabilistic in nature, this limits their appropriateness for tasks that demand a high degree of critical reasoning. More broadly, these insights are important when considering the role of AI in evidence synthesis. While AI may have potential to enhance our ability to retrieve information and accelerate some aspects of the review process, it cannot yet substitute for the nuanced judgment and expert oversight that humans provide (de la Torre-López et al., 2023).

Another key limitation for the use of AI in literature searching and evidence synthesis applications is Intellectual Property, and specifically the copyright and licensing restrictions that prohibit the ingestion of copyrighted material into generative AI tools. Indeed, apart from literature searches and title and abstract screening, all other stages of the review process (including full text screening, data extraction and critical appraisal) requires the model to have access to the full text. The development of applications of AI for these stages of the review process will be limited unless copyright and licensing agreements evolve.

## Current and future initiatives of the working group

The KLS AI working group was established two years ago as an interest-based exploratory horizon scanning group. The group has provided an effective forum to explore the applications, risks, and limitations of AI adoption into KLS workflows and discussing what impact we foresee AI having on KLS services. Earlier this year the working group transitioned from an interest-based and exploratory way of working to a more strategic approach to exploring the rapid development of AI-assisted technologies for information retrieval and evidence review processes. To support this, the working group is developing the infrastructure and resources required to support a more formal identification, prioritisation and evaluation of AI use-cases for each team within KLS services. This exercise is intended to ensure the effective and justifiable prioritisation of AI use-cases, considering factors such as tool cost, accessibility, time-saving potential, availability of external evaluation data, ethical concerns, and copyright and licensing restrictions.

Due to the pace at which AI technologies are developing, and the increased interest throughout the organisation, a separate AI working group that specifically focuses on the application of AI in evidence review processes was established in October 2023, working closely with the KLS group. Members of this working group are from several teams across UKHSA who regularly conduct evidence reviews as part of their day-to-day job, further facilitating a more focused and strategic approach to identifying opportunities for AI adoption throughout the organisation.

The promising findings from initial testing for some information retrieval and evidence review tasks has also been the foundation for our growing collaboration on AI across the organisation. For instance, members of the KLS AI working group are now partnering with data scientists in the UKHSA Data Analytics and Surveillance team to develop an internally hosted data extraction model using natural language processing. The approach taken throughout the development of this model reflects the working group’s general method for AI-related work—collaborative, iterative, and grounded in experience-based learning. The development process has provided a valuable opportunity for data scientists and evidence specialists to learn from each other and co-create what we anticipate will be an impactful and useful tool. Alongside the development and training of the model, the creation of a comprehensive evaluation framework has been central to this project. The model’s effectiveness will be evaluated using a previously published rapid review, comparing model-generated extractions to a human-extracted benchmark. Key aspects of the evaluation framework include assessing the model’s accuracy and acceptability compared to a human reviewer. For instance, various error types will be examined to evaluate the model’s accuracy and acceptability, and reliability is being assessed by reproducing each output at least ten times, evaluating consistency in style, tone, and content across all extraction attempts. The model’s evaluation is expected to be finalised in the coming months, and if it achieves the necessary level of accuracy, the model will replace the first reviewer, with its extractions checked by a second human reviewer in line with KLS standard rapid review methods.

The KLS AI working group has also faced some challenges, including lack of time to devote to AI activities outside of the working group and experiencing information overload given the pace at which AI is developing. It should also be noted that there are still many areas of AI’s application in library and information work that we have not explored yet, such as acquisitions and collection management, metadata and resource discovery. One reason for this may be that membership has not always included representatives across all functions of KLS. Capacity issues, resistance due to lack of trust but also fear of job loss, and need for further guidance and policy are other factors that may impact wider engagement and implementation of AI.

## Discussion and conclusions

AI and machine learning are not new concepts, however, the rapid progression of AI-assisted technologies that aid in various tasks relevant to knowledge, evidence and library services, such as information retrieval and evidence review processes, appears to be accelerating at pace. The establishment of an AI working group within KLS early in 2022 provided the opportunity to begin navigating the AI landscape and provided a forum to reflect on the implications of AI for KLS services, including the potential benefits and risks. It has also enabled the development of a strategic approach to scoping, testing, and evaluating these technologies to support their eventual integration, if and when appropriate. Alongside this, the working group has served as an excellent forum for gaining familiarity with AI-related concepts, methodologies, and terminologies, as well as for learning from each other. It has also provided valuable opportunities for collaboration with colleagues across the organisation who are working on AI and with external stakeholders, enabling us to share learning, experiences, and approaches to testing, evaluating, and implementing these technologies.

The tests conducted by the working group have shown that AI has great potential for knowledge and library services to save time and resources, including for searching and evidence review processes. Testing of machine learning and generative AI for literature searching have demonstrated the potential for AI to enhance our capacity for information retrieval and reference management, and some of the tools tested, namely Citationchaser and Deduklick, have now been implemented into KLS services.

The results from testing machine learning and generative AI for evidence review processes have also shown promise in review stages such as title and abstract screening, full text screening and data extraction, where AI can act as one reviewer and a human as the second reviewer. These stages are typically time-consuming and labour intensive, and the use of AI technologies could potentially increase our efficacy by facilitating parallel working, while having quality assurance processes in place to check AI-generated outputs. However, at present, machine learning tools for title and abstract screening are the only tools that have been implemented into KLS workflows for evidence review. For full text screening and data extraction, the main limitations remain copyright restriction which prevent further development of AI-assisted tools.

Moving forward, the working group will continue to address important ethical questions surrounding bias, transparency and the potential degradation of trust among users of AI-assisted evidence outputs. These concerns are highlighted in an Artificial Intelligence Health Security Threat Assessment conducted within UKHSA which emphasises the need for rigorous testing and transparent practices to mitigate risks such as bias and health inequalities, erosion of trust and the spread of mis-and disinformation (UKHSA, 2024). These issues are particularly present with generative AI, where the content of the training data used may lack transparency, unlike some types of machine learning, where the user can directly control training data. To mitigate these issues, the working group’s approach will need to incorporate transparent practices, such as ensuring model ‘explainability’ for any tools that KLS may adopt. This could involve adopting standards for documenting algorithm processes and educating users about the strengths and limitations of AI, including educating users to critically engage with and question AI-assisted outputs. This approach aligns closely with the organisational strategy, which advocates for a structured framework to ensure the ethical use of AI and emphasises the need for an ongoing assessment of AI’s impact in line with our strategic objectives and health security goals (UKHSA, 2024; UKHSA, 2023b).

Taking into account these considerations, rapid or systematic reviews are ideal use cases to implement AI-assisted tools while mitigating risks as each stage of the review is usually done entirely or partially in duplicate by two reviewers. Therefore, our approach to AI is to have AI acting as one reviewer, and the human acting as a second reviewer, ensuring adequate quality assurance processes and reduction of risk of bias that could be introduced by AI. On the other hand, the use of AI as one reviewer can also help reduce the risk of bias introduced by a human reviewer. Therefore, we envisage that the combination of both AI and human reviewers can potentially result in more reliable outputs while gaining efficiency. However, it is important to ensure adequate governance and processes are in place to control the use of AI and ensure we continue delivering the best available evidence for public health in a transparent and trustworthy manner.

In conclusion, by establishing an AI working group early in 2022, KLS has developed a thorough understanding of AI, its potential and its limitations within its activities and, more generally, within a public health organisation. This has allowed KLS to implement AI tools to increase the reliability and efficiency of its service, but also to support colleagues across the organisation in identifying potential use cases and conducting testing of AI tools.

## Data Availability

The data used in this paper is available from the corresponding author.

## Acknowledgements

The authors would like to thank all current and previous members of the Knowledge and Library Services (KLS) AI working group for their collective insights, which have contributed to this paper, including Alyson Foy, Anh Tran, Caroline De Brun, Edith Speller, Jennifer Hill, Jeremy Evans, Kirsty Morrison, Nicola Pearce-Smith, Rachel Gledhill, Sophie Pattison and Stephanie Grey. We also would like to thank Amy Sanders and Matthew Bosworth from the Science Evidence Review team (KLS, UKHSA) for their work on machine learning assisted screening, Harry Woolnough, Jonathan Fuller and Lauren Dunn from the Data Science Delivery Unit in UKHSA with whom KLS has been collaborating on various AI-related projects, and Carolina Arevalo for reviewing the manuscript.

## Availability of data and materials

The data used in this paper is available from the corresponding author.

## Author bibliographies

**Zalaya Simmons** is the Senior Evidence Reviewer at the UK Health Security Agency, within the Research, Evidence & Knowledge Division of the Scientific Strategy & Development Directorate. She holds an MSc in Health Policy, Planning, and Financing from The London School of Economics and Political Science and The London School of Hygiene and Tropical Medicine. Zalaya has years of experience in evidence review, contributing to diverse review topics ranging from infectious disease control to environmental health, resulting in various governmental publications. Her research interests include systematic review methods and the integration of machine learning and artificial intelligence in evidence review processes. She chairs the Knowledge and Library Services Machine Learning and AI working group at UKHSA.

**Charlotte Bruce** is a Knowledge and Evidence Specialist at the UK Health Security Agency within the Research, Evidence & Knowledge Division of the Scientific Strategy & Development Directorate. She holds a Masters in Information and Library Studies (Distinction) from Aberystwyth University. Charlotte has extensive experience in advanced public health evidence retrieval and is keen to investigate where AI can streamline service delivery and support the needs of her users. She is a founding member of the KLS Machine Learning and AI working group at UKHSA.

**Samuel Thomas** is the Knowledge and Library Services Manager at University Hospitals Dorset NHS Foundation Trust. He has previously worked at the UK Health Security Agency and Bournemouth University, and has a clinical background as an NHS Orthoptist. His professional interests include the exploration and implementation of AI and associated technologies in health libraries. He’s a committee member of the IFLA Artificial Intelligence Special Interest Group. Whilst working as a Knowledge and Evidence Specialist at UKHSA, Samuel set up and was the first Chair of the UKHSA KLS Machine Learning and AI Working Group.

**Patricia Lacey** is the Senior Knowledge and Evidence Manager for Local Authority Public Health, at the UK Health Security Agency within the Research, Evidence & Knowledge Division of the Scientific Strategy & Development Directorate. Trish has an MSc in library and information management and extensive experience in advanced public health evidence retrieval. Trish was chair of the AI working group in 2023 and her research interests include the use of AI within mis and disinformation.

**Wendy Marsh** is Head of Knowledge and Evidence Services at the UK Health Security Agency within the Research, Evidence & Knowledge Division of the Scientific Strategy & Development Directorate. Wendy has many years of experience in leading and delivering public health evidence services and currently heads a team that provides expert mediated access to the public health evidence base for UKHSA staff and partner organisations, delivers evidence reviews and leads UKHSA wide work on knowledge management and mobilisation.

**Scott Rosenberg** is the Head of Knowledge and Library Services at the UK Health Security Agency, within the Research, Evidence and Knowledge Division. Scott has an MSc in Healthcare Leadership, an NHS Leadership Academy Award in Senior Healthcare Leadership and a BSc in Information and Library Studies. A values led, ethical leader, Scott is a chartered member of CILIP and has significant experience of knowledge and information management and 14 years as a senior professional librarian in the health sector. Scott has strategic responsibility for library services, including three site libraries, library systems and evidence resource provision.

**Daphne Duval** leads the Science Evidence Review team within the Research, Evidence & Knowledge Division at the UK Health Security Agency. Daphne has extensive experience in producing evidence reviews to inform guidance, policy and research priorities in public health organisations. Daphne’s interests include rapid review methodologies for public health, use of evidence gap map to inform research priorities, and use of AI in evidence synthesis.

